# Attitudes Toward Prenatal Interventions in the Fanconi Anemia Community

**DOI:** 10.1101/2025.08.15.25333795

**Authors:** Tony Lum, Catherine Lee, Tippi MacKenzie, Billie Lianoglou, Agnieszka Czechowicz

## Abstract

**Background:** *In-utero* cell and gene therapies could become effective treatments for many inherited diseases. Notably, recent pre-clinical data supports that *in-utero* hematopoietic stem cell transplantation (*IU*-HSCT) can be curative for bone marrow disease in Fanconi anemia (FA) enabling preventative treatment without use of genotoxic conditioning agents or immune suppression. Given this, we surveyed patient and caregivers attitudes in the FA community toward prenatal diagnosis, treatment options, and clinical trials.

**Methods:** A multidisciplinary team created a comprehensive survey that was electronically distributed to the FA community by the leading FA patient advocacy group. Respondent’s demographic, history, and treatment information was collected and their attitudes toward *in-utero* therapies as well as termination of FA pregnancy was analyzed using univariable ordinal logistic regression

**Results:** 72 members from 18 countries completed the survey. 76% of respondents were willing to undergo *IU-*HSCT if FDA-approved while 68% were willing to enroll in a clinical trial to assess safety and efficacy of *IU-*HSCT or *IU-*gene therapy. 71% would undergo invasive testing to obtain a prenatal diagnosis of FA and 56% would be unlikely to end a pregnancy affected by FA.

**Conclusion:** *In-utero* therapy is a developing alternate treatment strategy for morbid inherited diseases. The FA community responds favorably to prenatal diagnosis and therapies, encouraging efforts to advance *IU*-cell and gene therapy clinical trials. This work may also provide insights into attitudes of other rare inherited disease communities.

## Background

There has been an explosion in the development of cell and gene therapies for many inherited diseases with approved treatments correcting diverse cell-types from hematopoietic cells such as in sickle cell anemia (CASGEVY®, LYFGENIA^TM^) to neurologic cells such as in spinal muscular atrophy (ZOLGENSMA®). While these therapies are currently utilized in the postnatal setting, prenatal intervention could enable preventative treatment for many inherited diseases before birth. However, it is unknown how patients and caregivers would respond to the availability of such therapies.

Fanconi anemia (FA) is a rare genetic disorder that is representative of many genetic diseases, causing morbidity in patients at various stages of life which often starts in early childhood. To date, 23 genes have been implicated in FA, with inheritance patterns ranging from X-linked recessive to autosomal dominant, though most cases are autosomal recessive(Gille et al., 2012). FA is characterized by an impaired ability to repair intra-strand crosslinks, leading to accumulating DNA damage(Soulier, 2011) Kitao & Takata, 2011). Approximately 90% of patients with FA will ultimately experience bone marrow failure, resulting in severe multi-lineage cytopenia that which requires persistent hematopoietic support(Kutler et al., 2003; Soulier, 2011). Optimally this could be provided through a one-time curative cell or gene therapy treatment.

Postnatal allogeneic (allo) hematopoietic stem cell transplantation (HSCT) from a healthy donor is currently the standard of care for FA-related hematopoietic disease (Supplemental Figure 1)(Dufour & Pierri, n.d.). However, the procedure is time and resource-intensive, has high costs, and is associated with significant morbidity. The current allo-HSCT process involves identifying and collecting hematopoietic cells from a suitable donor, conditioning the patient with chemotherapy and/or irradiation to both clear space for donor stem cell engraftment and prevent immunologic rejection, and a 4-8 week hospitalization for the transplant and immediate recovery. Post-transplant recovery also requires an additional period of isolation to mitigate risks associated with a recovering immune system. Additional complications often include mucositis, organ damage, and graft-versus-host disease (GVHD). Furthermore, most FA patients eventually develop secondary cancers due to the genotoxic effects of the chemoradiotherapy given their inability to repair DNA damage(Alter et al., 2018; Dufour & Pierri, n.d.; Mathew, 2006). Consequently, FA patients qualify for allo-HSCT only when they develop severe hematopoietic disease due to bone marrow failure or hematologic malignancy (Mathew, 2006). The same is true for many hematolymphoid diseases, which are currently treated only once patients develop advanced symptoms due to current challenges and risks of HSCT.

Our team and others have been pioneering alternative treatment options for rare inherited diseases that could be used in the prenatal setting to improve treatment outcomes. Notably, we have developed both *in-utero hematopoietic stem cell transplantation* (*IU-*HSCT) as well as *in-utero* enzyme replacement therapies (*IU*-ERT)(Cohen et al., 2022; Schwab et al., 2023; Tippi C. MacKenzie, 2017; Witt et al., 2018), with future potential for *in-utero* gene therapy (*IU*-GT). Notably, FA is a unique disease where the hematopoietic system has the potential to be cured prenatally through *IU-*HSCT alone without the need for supplemental drugs or therapies. Specifically, preclinical studies in various FA mouse models from our group and a complementary group suggest that *IU*-HSCT could be an effective therapeutic approach for the bone marrow failure caused by this condition without any toxicity (Swartzrock et al., 2024; Nijagal et al., 2013) (Supplemental Figure 2). In this scenario, fetuses diagnosed prenatally with FA would be prophylactically treated using the mother as the hematopoietic stem cell (HSC) donor. The fetus is able to accept maternal HSCs without conditioning agents due to its unique immune system that is primed for tolerance(Cowan et al., 2001). This minimally invasive procedure would be performed in the second trimester, with maternal HSCs transplanted via an ultrasound-guided intravenous injection into the fetal umbilical vein. If translatable, *IU*-HSCT could prevent future bone marrow failure while avoiding the current toxicities associated with postnatal allo-HSCT. This strategy parallels the *ex-vivo* FANCA lentiviral gene therapy that is in development, which aims to treat FA patients early to prevent hematologic disease without chemoradiotherapy. Though there are limitations to this gene therapy including its exclusive use in patients with FANCA mutations, high resource demands, incomplete efficacy, and risks associated with lentiviral integration such as clonal hematopoiesis or malignancy(Czechowicz et al., 2022; Río et al., 2019). *IU*-HSCT holds the potential to overcome each of these limitations, creating a new preventative treatment paradigm for inherited genetic diseases.

While *IU-*HSCT represents a novel treatment option with the potential to mitigate the risks and side effects associated with current therapies for FA, perspectives of patients and caregivers are critical to the development and implementation of any new treatment. Thus, we designed and disseminated a survey to the FA community that provided background regarding the *IU*-HSCT approach and how it compares to the current treatments (Supplemental Figures 2 and 1, respectively). Using this survey, we ascertained the attitudes towards prenatal diagnosis, termination and *IU-*HSCT. Further, we also determined attitudes towards *IU-*GT which could enable systemic disease correction and eliminate need for donor cells. These attitudes may parallel those of other inherited disease patient communities which are also important to consider as various prenatal interventions are being developed. The results of this survey of the FA community are presented here.

**Figure 1:**
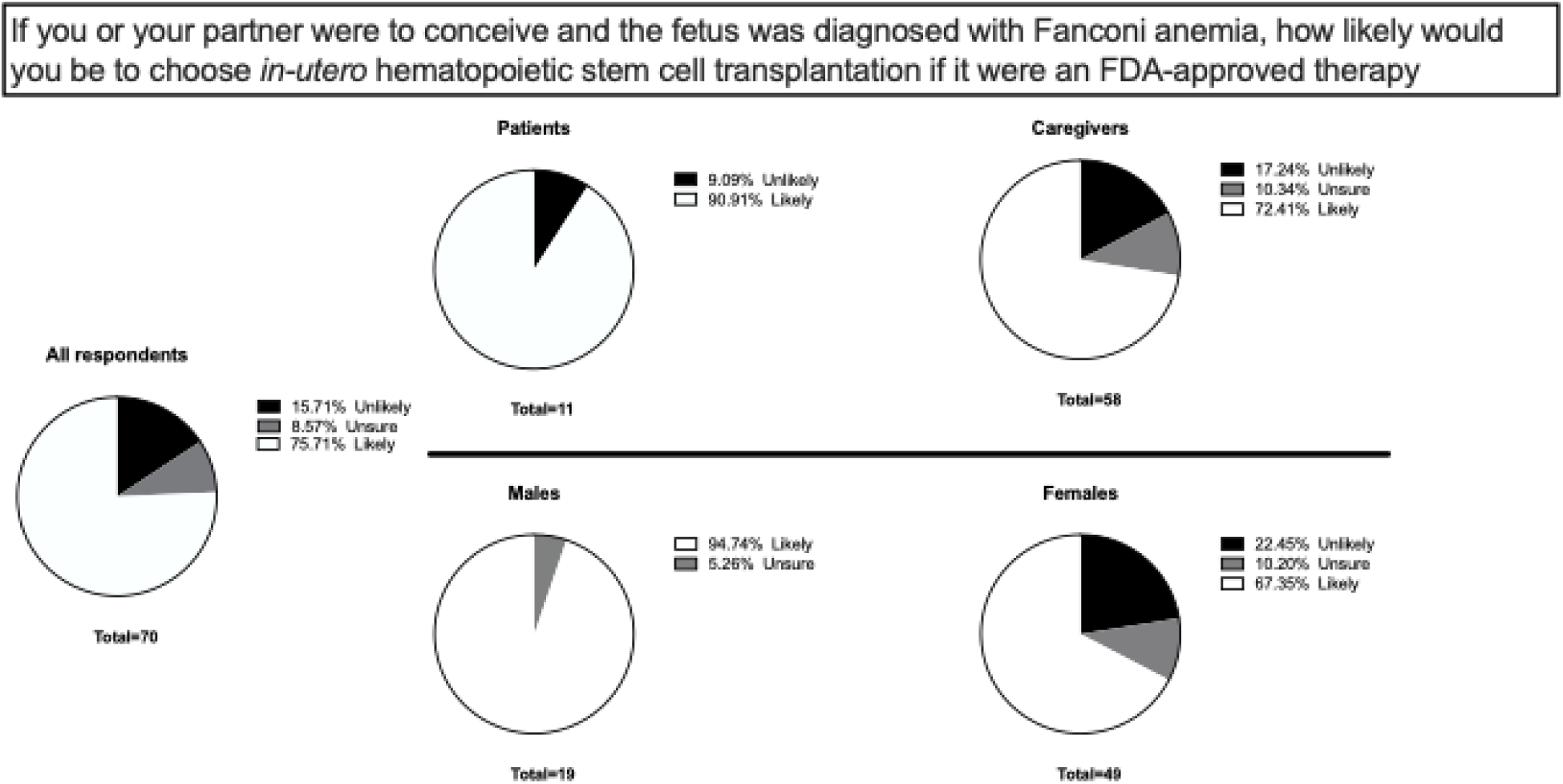
Attitude toward acceptance of an FDA approved *in-utero* hematopoietic stem cell transplantation in the indicated responder groups. Respondents were asked “If you or your partner were to conceive and the fetus was diagnosed with Fanconi anemia, how likely would you be to choose *in-utero* hematopoietic stem cell transplantation if it were an FDA-approved therapy?”. Comparing acceptance of an FDA-approved *in-utero* hematopoietic stem cell transplantation between male and female respondents, Fisher’s exact test: (OR 9.16, 95% CI 1.12, 74.6, p=0.04).

## Methods

### Instrument Development

The goal of this study was to assess the attitudes of FA patients and caregivers toward an *in-utero* treatment approach. No validated questionnaire was available on this topic, so our team created an electronic questionnaire that was reviewed by a multidisciplinary team (genetic counselor, physicians, and patient advocacy team members). Questions were created and modified in a structured response format with a free text option where appropriate. The survey contained 22 questions. After the respondent categorized themselves into patient or caregiver, the survey branched into 3 sections. The first section asked about specific diagnostic details and therapies received by the affected individual; the second section evaluated attitudes regarding termination of pregnancies affected by FA, *in-utero* treatment options, and clinical trials using a 5-point Likert scale; the third section surveyed respondent demographic characteristics, educational sources, and healthcare coverage of the patient. Healthcare choices that included “Medicaid” and “other” were consolidated into “federally funded insurance” once analysis of free text answers revealed healthcare was provided by respondent’s respective government.

### Survey Distribution

The survey was approved by the University of California, San Francisco Institutional Review Board (IRB#: 19-27109). A link to the e-survey, created on REDCap, was distributed to the FA community by the Fanconi Cancer Foundation ([FCF]; previously known as the Fanconi Anemia Research Fund [FARF]). FCF is a nonprofit organization and leading FA patient advocacy group in the United States with global outreach. FCF provided the FA infographic and survey link in monthly newsletters. The survey was anonymous and remained available from 6/4/2024 until 9/24/24.

### Statistical Analysis

Duplicate and empty surveys were excluded from analysis. Descriptive statistics and Fisher’s Exact tests were used to assess the relationship between the categorical outcome variables and respondent characteristics (caregivers vs. patients and males vs. females). For the outcome variables measured using a 5-point Likert scale, we collapsed “extremely unlikely” and “somewhat unlikely” and “somewhat likely and “extremely likely”. Univariable ordinal logistic regression was used to analyze associations between respondent characteristics and the following outcome variables: (1) choose *IU-*HSCT if it were an approved therapy, (2) enroll in a clinical trial to determine safety and efficacy for *IU-*HSCT, (3) enroll in a clinical trial to determine safety and efficacy for *IU-*gene therapy, and (4) ending a pregnancy affected by FA. All statistical analyses were performed using R Statistical Software (version 4.4.1 R Core Team 2024).

## Results

### Respondent Characteristics

An online survey was developed by a multidisciplinary team and distributed to the FA community (Supplemental Figure 3). A total of 72 individuals responded, made up of 58 caregivers (81%), 12 patients (17%), and 2 unreported (3%). The majority of respondents were white (90%), from the United States (67%), and female (71%) (Table 1). 61% of respondents reported the patient having employer-based health insurance, 25% were federally insured, and the remainder independently purchased their insurance or were unreported (Table 2). Among the caregivers who answered, 40 had one child with FA (74%), 11 had children with and without FA (20%), and 3 reported having multiple children with the condition (6%). The most common gene variant identified among respondents was FANCA (51%) (Tables 1 and 2).

**Table 1:** Demographics of respondents (n=72) including their race and location.

| **Respondent Characteristics** | **N (%) or median (range)** |
| --- | --- |
| **Demographics** |  |
| Caregivers | 58 (81%) |
| *One child with FA* | 40 (74%) |
| *Multiple children with FA* | 3 (6%) |
| *Children with and without FA* | 11 (20%) |
| Patients | 12 (17%) |
| *Unreported* | 2 (3%) |
| Age (years) | 48 (26-82) |
| **Sex** |  |
| Female | 51 (71%) |
| Male | 19 (26%) |
| *Unreported* | 2 (3%) |
| **Race** |  |
| American Indian/Alaskan Native | 1 (1%) |
| Asian | 2 (3%) |
| Hispanic or Latino | 1 (1%) |
| Native Hawaiian/Pacific Islander | 1 (1%) |
| White | 65 (90%) |
| Multiracial or biracial | 1 (1%) |
| Other | 1 (1%) |
| **Geographic Location** |  |
| USA | 48 (67%) |
| Europe | 16 (22%) |
| Canada | 2 (3%) |
| Australia | 2 (3%) |
| Africa | 2 (3%) |
| S America | 1 (1%) |
| *Unreported* | 1 (1%) |

**Table 2:**
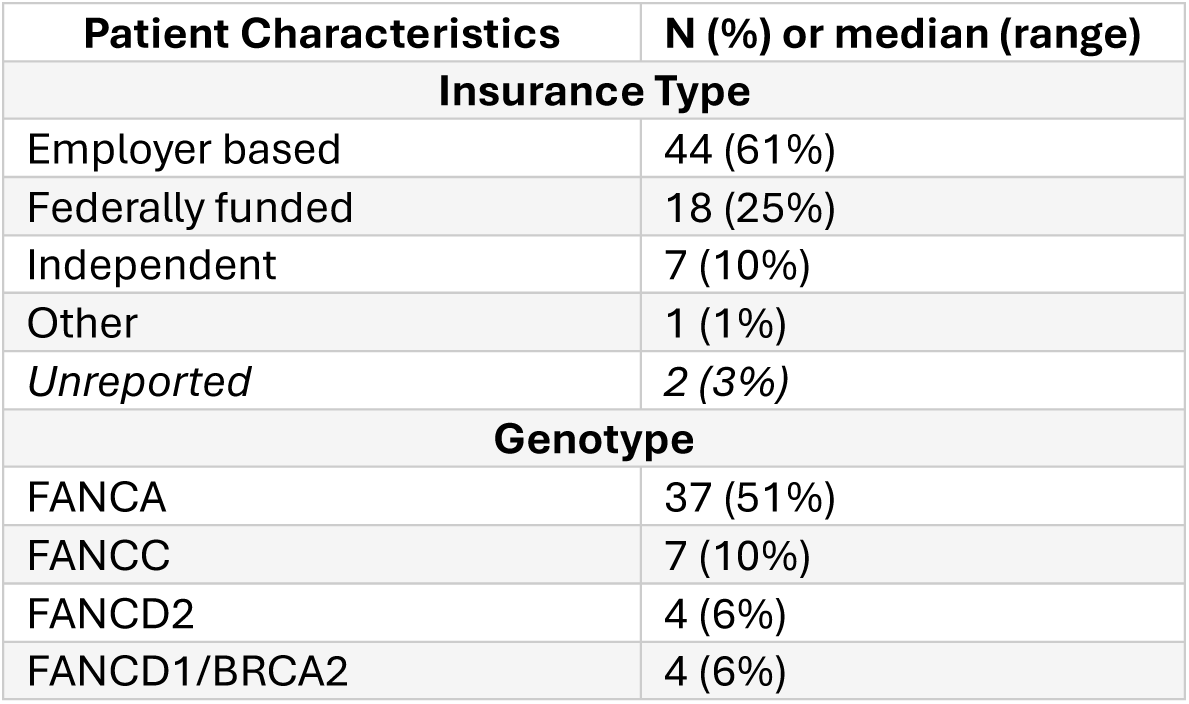
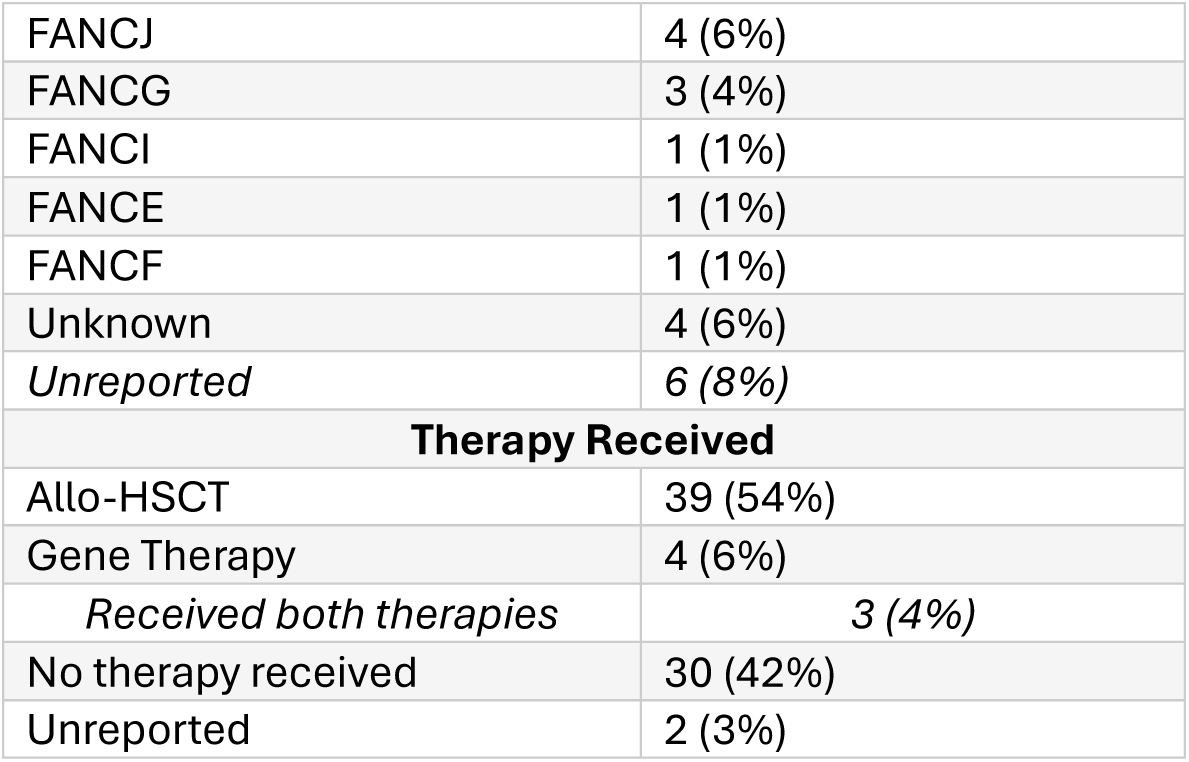
Respondents listed health insurance, genotype, and therapy received for Fanconi anemia patients.

### Respondent Attitudes Toward Prenatal Therapies and Clinical Trials

As we develop treatment options for FA, it remains important to survey the community’s acceptance. This is especially important for paradigm shifting prenatal therapy approaches. Overall, 76% of respondents expressed willingness to undergo *IU-*HSCT if it were FDA-approved (Figure 1). Demographic analysis revealed that both FA patients (91%) and FA caregivers (72%) indicated they would likely pursue *IU-*HSCT if it were an approved treatment option (Figure 1). Notably, males were particularly open to this treatment option, with 95% in favor compared to 67% of females (OR 9.16, 95% CI 1.12, 74.6, p=0.04) (Figure 1).

Clinical trials are necessary for the development of new therapies and are a critical component of assessing the safety and efficacy of new treatments. When asked about their willingness to enroll in clinical trials for *IU-*HSCT or *IU*-GT, 68% of respondents were open to participation to either treatment (Figure 2). Male respondents were again the most likely to enroll with 84% expressing willingness, compared to 58-60% of females (Figure 2).

**Figure 2:**
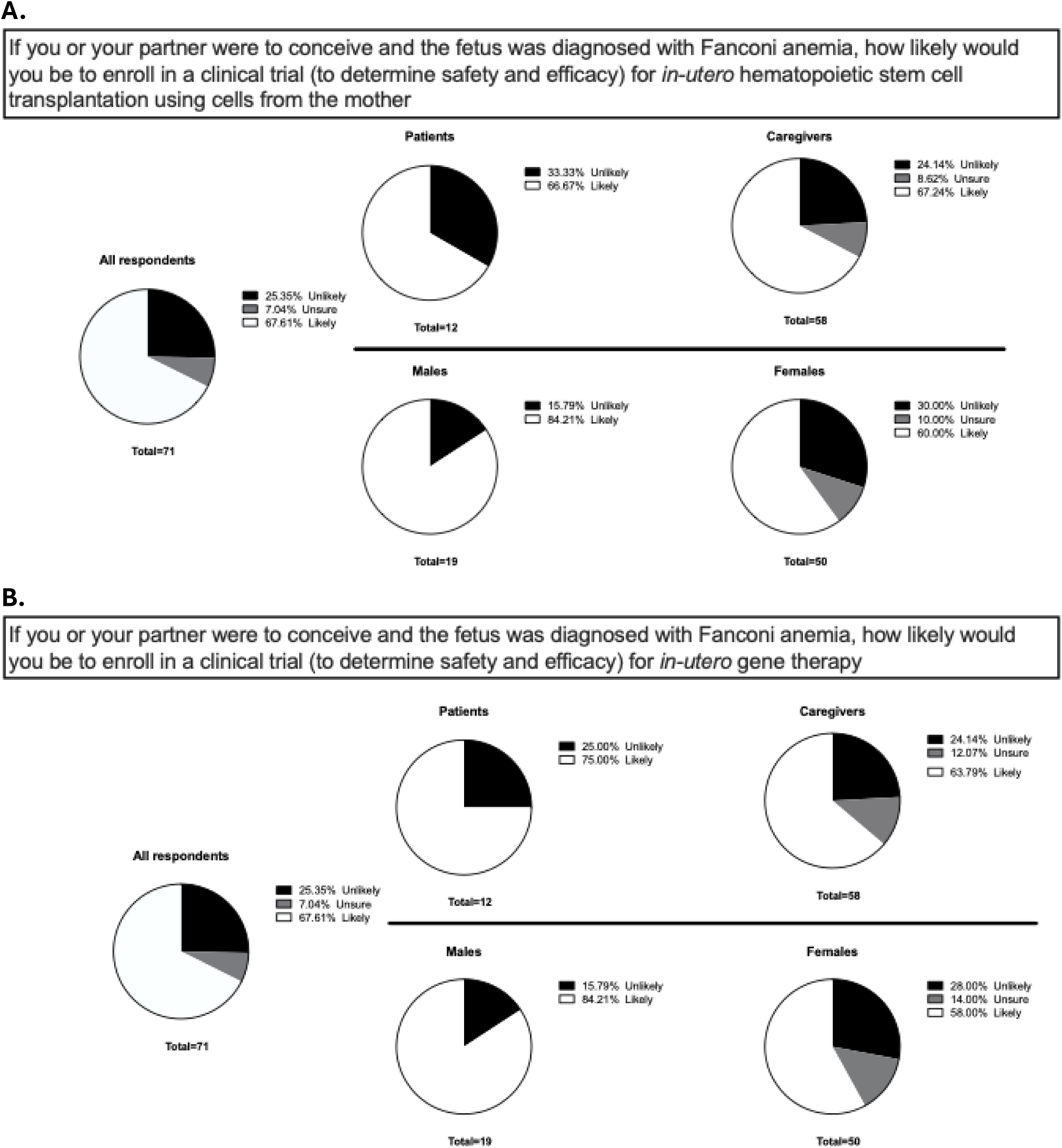
Attitudes toward enrolling in a clinical trial for *in-utero* hematopoietic stem cell transplantation (**A**) or *in-utero* gene therapy (**B**) in the indicated responder groups. Respondents were asked “If you or your partner were to conceive and the fetus was diagnosed with Fanconi anemia, how likely would you be to enroll in a clinical trial (to determine safety and efficacy) for *in-utero* hematopoietic stem cell transplantation using cells from the mother or *in-utero* gene therapy?”

### Respondent Attitudes Toward Prenatal FA Diagnosis and Termination

As prenatal diagnosis is critical for prenatal treatment, survey participants were also asked about their views on pursuing prenatal diagnostic procedures. Currently, FA is not part of routine prenatal screening and suspicion of disease often occurs due to a positive family history or abnormalities detected on ultrasound. Diagnostic confirmation is available through invasive procedures such as amniocentesis or chorionic villus sampling, methods which carry small risks of miscarriage and/or infection. Despite these risks, 71% of all respondents stated they would pursue prenatal diagnosis in any pregnancy with 100% of patients in agreement (Figure 3).

**Figure 3:**
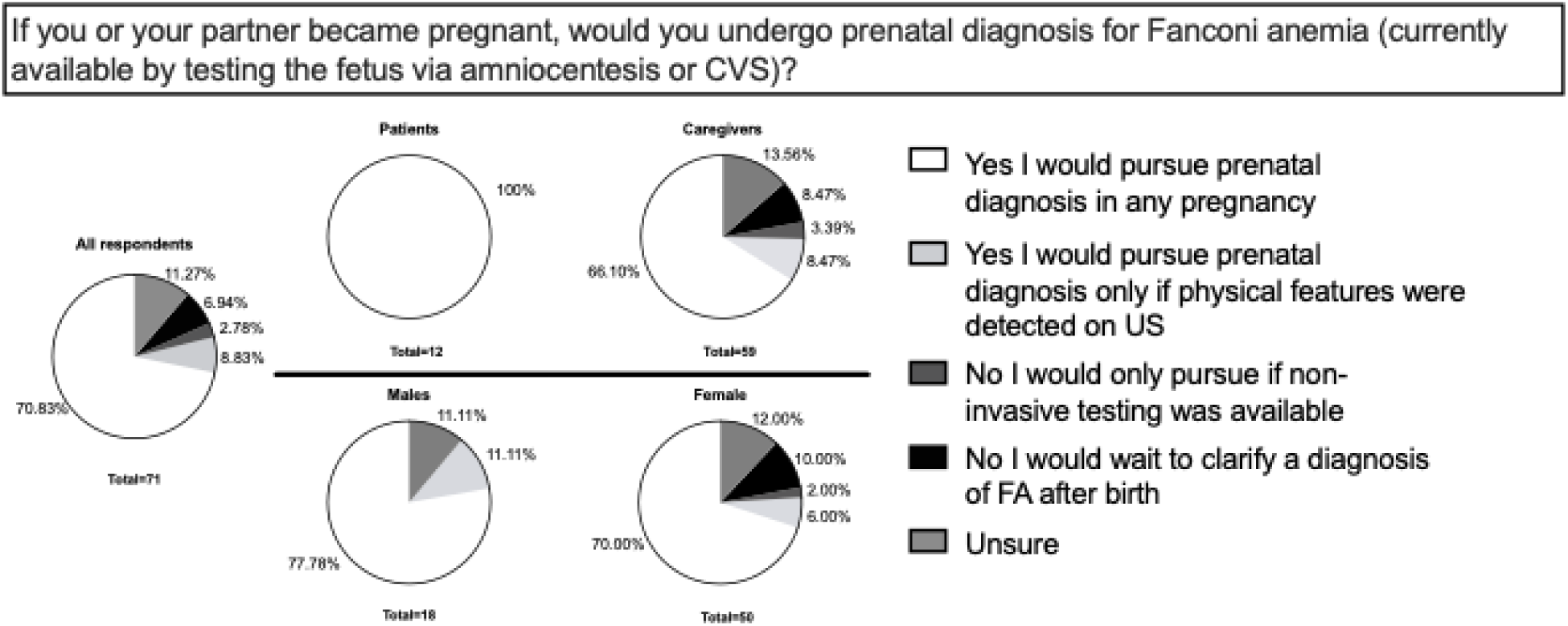
Attitudes toward invasive prenatal diagnostic testing for Fanconi anemia in the indicated responder groups. Respondents were asked “if you or your partner became pregnant, would you undergo prenatal diagnosis for Fanconi anemia (currently available by testing the fetus via amniocentesis or CVS)?”. Comparison of answers between male and female respondents as well as patient and caregiver respondents.

Given the variability in FA severity, pregnancy decisions can be especially challenging. When asked about the possibility of terminating a pregnancy affected by FA, approximately half of respondents indicated they would consider this option (Figure 4). Respondents with employer-based insurance were more likely to consider termination compared to those with federally-funded or independently purchased insurance (OR 4.6, 95% CI 1.31, 16.07, p=0.02 and OR 7.46, 95% CI 0.83, 67.29, p=0.07; respectively) (Figure 4).

**Figure 4:**
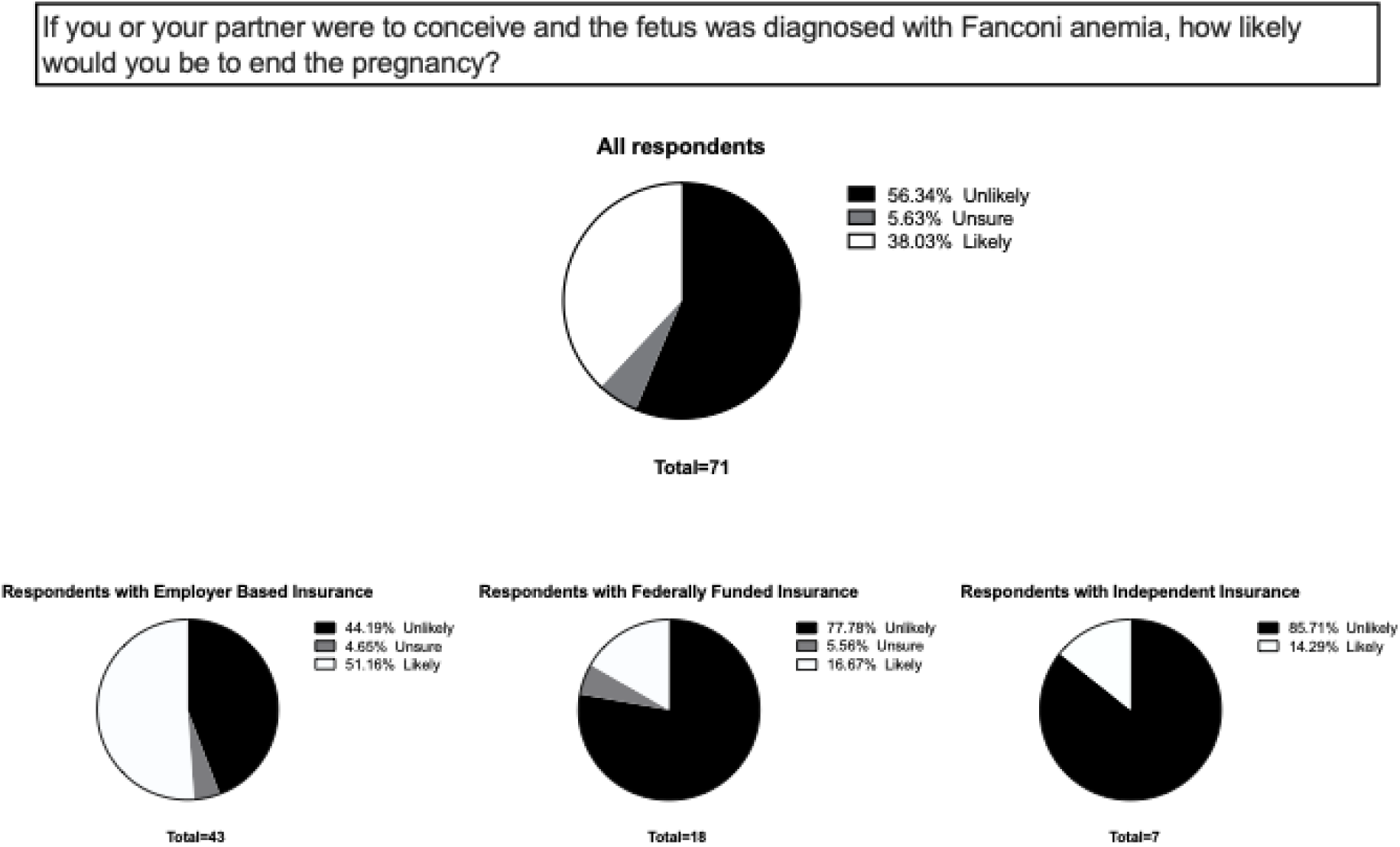
Attitude toward ending a Fanconi anemia affected pregnancy in the indicated responder groups. Respondents were asked “If you or your partner were to conceive and the fetus was diagnosed with Fanconi anemia, how likely would you be to end the pregnancy?” Comparing likelihood of ending pregnancy between patients with employer based and federally funded insurance types, Fisher’s exact test: (OR 4.6, 95% CI 1.3, 16.1, p=0.02).

## Discussion

Given the potential for *IU-*HSCT in FA patients to become a near-term, preventative therapy which shifts the treatment paradigm for this challenging inherited disease, we conducted a comprehensive survey on the attitudes on the FA community toward prenatal diagnosis, treatment options, and cell and gene therapies. In our survey of FA patients and caregivers, we found that most respondents were amenable to an *in-utero* treatment aimed at preventing future bone marrow failure associated with this congenital disease. Additionally, the majority expressed willingness to enroll in clinical trials for *IU-*HSCT or *IU-*GT. Notably, male respondents were more likely to opt for prenatal diagnostic procedures and/or interventions when compared to females. This may in part be due to the presented intervention being carried out on the mother, and attitudes might be altered if non-invasive diagnostic testing or paternal HSC donation were possible.

FA is a rare disease, with an estimated incidence rate of 1 in 136,000(Che et al., 2018). It is currently not included in standard prenatal or postnatal screening protocols, and testing is typically initiated only after the detection of an abnormal finding or its presence in a family history. At present, FA prenatal diagnosis is only available through invasive procedures, such as amniocentesis or chorionic villus sampling, which all patient respondents reported they would pursue despite associated risks(*American College of Obstetricians and Gynecologists*, 2025; Carlson & Vora, 2017). While most respondents indicated they would continue a pregnancy affected by FA, a significant proportion would consider termination with higher likelihood in those with employer-based insurance. This decision is presumably influenced by the severe morbidity presently associated with FA as well as the costly, complex, and high-risk current treatment options.

It remains critical to amplify the voices of patients and caregivers when developing and considering new experimental treatments. *IU-*HSCT is a procedure with ethical implications as it affects two patients—the mother and the fetus. Deciding whether to enroll in a clinical trial for such treatments involves weighing the certainty of a congenital disease against the uncertainty of experimental therapy outcomes, including potential unknown side effects and lack of guaranteed efficacy. Despite this ethical dilemma, our findings suggest that individuals affected by FA are generally supportive of innovative approaches like *IU-*HSCT, which could offer those affected a chance for an equitable start to life. These attitudes may parallel those of other inherited disease patient communities and are important to consider when counseling various patients and developing new treatment approaches.

### Strengths and Limitations

This study has notable strengths. It represents one of the first efforts to gauge the attitudes of the FA community toward novel *in-utero* therapies. By focusing on a rare disease population, this survey provides valuable insights into a community often underrepresented in discussions about experimental treatments. Diverse representation of respondents was observed with participation across races, geographical areas, insurance types and genotypes. The use of infographics to compare and contrast treatment options may also help respondents make more informed decisions.

However, the study also has limitations. There were limited number of males who responded and responders were surveyed individually rather than as couples, so it is unclear how generalizable these answers would be to the community where dual parental consent is often required. Also, the willingness to choose an *in-utero* treatment option or enroll in a clinical trial may not translate into actual participation. In addition, the survey was distributed electronically through a patient care organization, meaning that the respondents represent only a subset of those affected by FA. Furthermore, the reliance on electronic distribution may have excluded individuals without internet access, which could disproportionally affect underrepresented and underserved individuals in healthcare research. This limitation was evident as over 90% of respondents were from Western countries.

## Conclusion

Our findings highlight support within the FA community for *in-utero* therapies including *IU-* HSCT and *IU*-GT. Given *IU-*HSCT has demonstrated clinical safety and feasibly for other diseases and given recent pre-clinical data illustrates robust safety and efficacy of this approach in multiple FA mouse models, *IU-*HSCT in FA has the potential to become a near-term preventative clinical treatment for patients with this disease laying a new paradigm for treatment of inherited diseases. While additional steps remain in transforming this support into an *IU-*HSCT clinical trial for FA patients, the willingness of patients and caregivers to consider these treatments emphasizes the importance of continued research and innovation. Future efforts should focus on addressing barriers to trial participation, ensuring equitable access, improving prenatal diagnosis and diagnostics, and continual incorporation of patient and caregiver perspectives into development of clinical trials involving new therapeutic options. Moreover, additional research into the attitudes of other patient communities is greatly needed as new therapies for these diseases are also being developed.

## Data Availability

All data produced in the present study are available upon reasonable request to the authors

## List of abbreviations and acronyms

Allo: allogeneic
ERT: enzyme replacement therapy
FA: Fanconi anemia
GT: gene therapy
GVHD: graft-versus-host disease
HSC: hematopoietic stem cell
HSCT: hematopoietic stem cell transplantation
IU: in-utero

## Declarations

### Disclosures

A.C. discloses financial interests in the following entities working on gene-therapy and antibody-based conditioning approaches: Beam Therapeutics, Editas Medicines, GV, Inograft Biotherapeutics, and Prime Medicines. In addition, she is an inventor on antibody-based conditioning patents licensed to Jasper Therapeutics, Gilead Sciences, Inograft Biotherapeutics and Magenta Therapeutics and has received sponsored research funding from Jasper Therapeutics, Rocket Pharma, and STRM.Bio. T.C.M. has received grant funding from companies in the rare disease space, including Novartis, BioMarin, Biogen, and Ultragenyx. The remaining authors declare no competing financial interests.

### Authors’ Contributions

T.L.: Designed the study, curated and analyzed data, contributed to writing-original draft, writing-review and editing. C.L.: Analyzed data, contributed to writing-review and editing. T.M.: Conceptualized the study, provided resources, mentorship and supervision, contributed to writing-review and editing. B.L.: Conceptualized and designed the study, provided resources, mentorship and supervision, contributed to writing-review and editing. A.C.: Conceptualized and designed the study, acquired funding and support, provided resources, mentorship and supervision, contributed to writing-review and editing.

## Acknowledgments

Special thanks to the FA patients and community that was critical in the study design and study conduct, with notable contributions from the FCF leadership team including Isis Sroka, Jordan Deines, and Taylor Schreiner who made this study possible. The authors would also like to thank Drs. Yair Blumenfeld, Yasser El-Sayed, Anna Girsen, and the Stanford University School of Medicine Dunlevie Maternal-Fetal Medicine Center for Discovery, Innovation and Clinical Impact for their support of this work, as well as Ms. Molly McGuinness for added input and support.

## Funding

This study was supported by a grant to A.C. from the Fanconi Cancer Foundation (formerly Fanconi Anemia Research Fund).

## Ethics approval and consent to participate

The survey was approved by the University of California, San Francisco Institutional Review Board (IRB#: 19-27109).

## Consent for publication

Not applicable.

## Availability of data and materials

All data generated or analyzed during the current study are included in this published article and its supplementary information files.

## Competing interests

The authors declare that they have no competing interests.

## Notes

### Competing Interest Statement

The authors have declared no competing interest.

### Author Declarations

The survey was approved by the University of California, San Francisco Institutional Review Board (IRB#: 19-27109) of the University of California, San Francisco gave ethical approval for this work

