## Supplementals Figures for "Attitudes Toward Prenatal Interventions in the Fanconi Anemia Community"

### Supplemental Figures

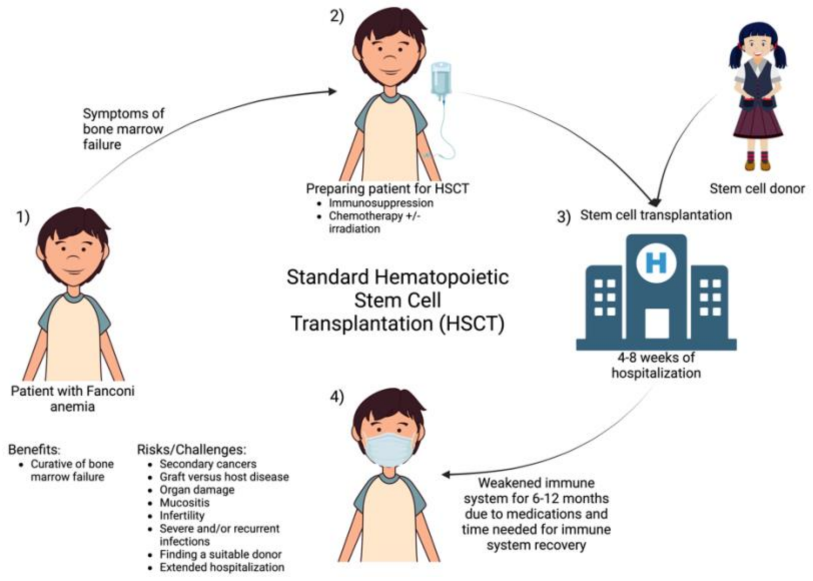


**Supplemental Figure 1**: Infographic created to educate respondents on current approved treatment: “Standard Hematopoietic Stem Cell Transplantation” for Fanconi Anemia patients experiencing bone marrow failure.

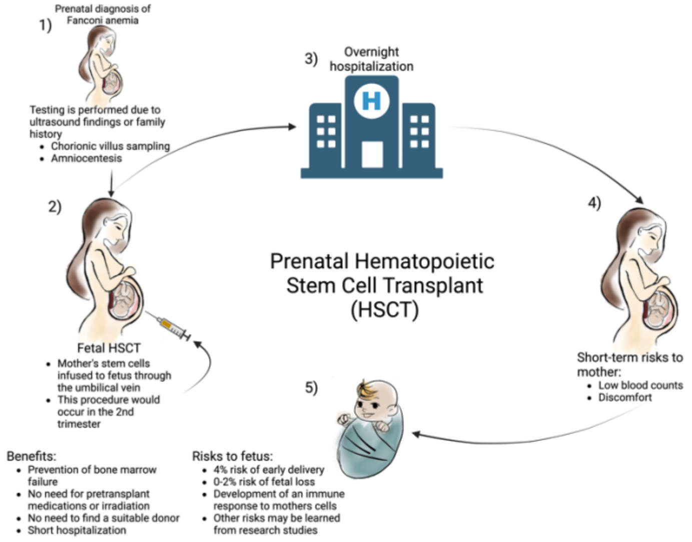


**Supplemental Figure 2:** Infographic created to educate respondents on proposed treatment: “Prenatal Hematopoietic Stem Cell Transplantation” to prevent possible future bone marrow failure in Fanconi Anemia patients.

**Questionnaire:**

I am a:

1) Patient with Fanconi anemia
2) Caregiver of a patient with Fanconi anemia

I have:

1) One child with Fanconi anemia
2) Multiple children with Fanconi anemia
3) Both children with and without Fanconi anemia

If known, what Fanconi anemia gene variant do you or your children carry? (e.g. which FANC gene is mutated or is this unknown)

1) Free text

At what ages was (were) you or your child(ren) diagnosed (check all that apply)?

1) Before birth
    a. Please specify gestational age (in weeks) and what prompted prenatal testing
2) After birth and before development of symptoms
    a. Please specify age (in years) and what prompted diagnostic testing
3) After birth and after development of symptoms
    a. Please specify age (in years) and symptoms

Has the patient received hematopoietic stem cell transplantation (HSCT)

1) Yes
2) No

Has the patient received gene therapy

1) Yes
    a. Please specify
2) No

If you or your partner became pregnant, would you undergo prenatal diagnosis for Fanconi anemia (currently available by testing the fetus via amniocentesis or chorionic villus sampling [CVS])

1) Yes, I would pursue prenatal diagnosis in any pregnancy
2) Yes, I would pursue prenatal diagnosis but only if physical features of Fanconi anemia were detected via ultrasound
3) No, I would only pursue testing in pregnancy if a non-invasive blood test was available (not currently possible)
4) No, I would wait to clarify a diagnosis of Fanconi anemia after birth
5) Not sure
    a. Please explain

**Likert Survey**: If you or your partner were to conceive and the fetus was diagnosed with Fanconi anemia prenatally, how likely would you be to:

| | Extremely Unlikely | Somewhat unlikely | Neither likely nor unlikely | Somewhat likely | Extremely likely |
|---|---|---|---|---|---|
| End the pregnancy | | | | | |
| Choose prenatal HSCT if it were an FDA approved therapy | | | | | |
| Enroll in a clinical trial (to determine safety and efficacy) for prenatal HSCT using cells from mother | | | | | |
| Enroll in a clinical trial (to determine safety and efficacy) for prenatal gene therapy | | | | | |
| Continue the pregnancy without fetal intervention and consider preventative gene therapy after birth (If available) | | | | | |
| Continue pregnancy without prenatal intervention and pursue HSCT if symptoms of bone marrow failure occur | | | | | |

What types of information would be helpful to better understand the prenatal transplant process
1) Free text

Please share any thoughts you have that were not specifically asked about in this survey
1) Free text

#### **Demographics:**

How old are you
1) Free text

What is your biological sex
1) Female
2) Male

Where do you get most of your information about treatment options (please select one answer)
1) Physicians
2) Patient organizations like Fanconi cancer foundation (formerly Fanconi Anemia Research Fund)
3) Clinicaltrials.gov
4) Other
    a. Please specify

What country do you live in (if in USA, what state)?
1) Free text

What is the primary type of health coverage the patient has?
1) Employer based
2) Medicaid
3) Independently purchased
4) Other
    a. Please specify

Which of the following best describes the patient (select all that apply)
1) American Indian or Alaskan native
2) Asian
3) Black or African American
4) Hispanic or Latino
5) Native Hawaiian or Pacific Islander
6) White or Caucasian
7) Multiracial or biracial
8) Other
    a. Please specify

**Supplemental Figure 3:** Questionnaire created by multidisciplinary team for patients and caregivers with Fanconi Anemia.
